# Evaluation of coronavirus decay in French coastal water and application to SARS-CoV-2 risk evaluation using Porcine Epidemic Diarrhea Virus as surrogate

**DOI:** 10.1101/2023.04.18.23288060

**Authors:** Maud Contrant, Lionel Bigault, Mathieu Andraud, Marion Desdouits, Sophie Rocq, Françoise S. Le Guyader, Yannick Blanchard

## Abstract

SARS-CoV-2 in infected patient mainly display pulmonary and oronasal tropism however, the presence of the virus has also been demonstrated in stools of patients and consequently in wastewater treatment plant effluents, questioning the potential risk of environmental contamination (such as seawater contamination) through inadequately treated wastewater spill-over into surface or coastal waters. The environmental detection of RNA alone does not substantiate risk of infection, and evidence of an effective transmission is not clear where empirical observations are lacking.

Therefore, here, we decided to experimentally evaluate the persistence and infectious capacity of the Porcine epidemic diarrhea virus (PEDv), considered as a coronavirus representative model and SARS-CoV-2 surrogate, in the coastal environment of France. Coastal seawater was collected, sterile-filtered, and inoculated with PEDv before incubation for 0–4 weeks at four temperatures representative of those measured along the French coasts throughout the year (4, 8, 15, and 24°C). The decay rate of PEDv was determined using mathematical modeling and was used to determine the half-life of the virus along the French coast in accordance with temperatures from 2000 to 2021.

We experimentally observed an inverse correlation between seawater temperature and the persistence of infectious viruses in seawater and confirm that the risk of transmission of infectious viruses from contaminated stool in wastewater to seawater during recreational practices is very limited. The present work represents a good model to assess the risk of transmission of not only SARS-CoV-2 but may also be used to model the risk of other coronaviruses, specifically enteric coronaviruses.

**Importance:** This present work is a follow up addressing the question of the persistence of coronavirus in marine environment owing to the fact that SARS-CoV-2 is regularly detected in wastewater treating plan and the coastal environment is particularly at risk since it is subjected to increasing anthropogenic pressure and is the final receiver of surface waters and treated or sometimes insufficiently depurated waste waters. Our findings are of interest to researchers and authorities seeking to monitor SARS-CoV-2 and also enteric coronaviruses in the environment, either in tourist areas or in regions of the world, where centralized systems for wastewater treatment are not implemented, and more broadly, to the scientific community involved in “One Health” approaches.

## Introduction

The emergence of the human coronavirus, SARS-CoV-2, accompanied by its worldwide spread leading to the COVID pandemic (671 million cases and 6.85 million deaths on February 2023) (WHO (World Health Organization), reminds us, if needed, the health hazard posed by coronaviruses.

Human coronaviruses (HCoVs) are respiratory viruses that are primarily transmitted by exposure to droplets generated by coughing, sneezing or breathing, either directly in the respiratory tract or indirectly through contact with surfaces contaminated by these droplets (Marques and Domingo, 2021; Zhang et al., 2020a). Interestingly, SARS-CoV-2 RNA has been repeatedly detected in stool samples of infected patients (Xiao et al., 2020; Zhang et al., 2020b), for review (Jones et al., 2020), for long periods (Wu et al., 2020) even in the absence of gastrointestinal symptoms (Han and He, 2021; Tang et al., 2020), questioning in the early time of the pandemic the potential risk of fecal-oral or fecal-respiratory transmission (Ahmed et al., 2020a; Arslan et al., 2020; Dona et al., 2020; Gu et al., 2020; Heller et al., 2020; Hindson, 2020; Shutler et al., 2021). The SARS-CoV-2 genome was detected in raw wastewater from different metropolitan areas including Paris (Wurtzer et al., 2020), with concentrations correlating with the estimated numbers of Covid-19 cases. The genome has also been detected in treated effluents from sewage treatment plants, but to a lesser extent, suggesting that SARS-CoV-2 may contaminate the environment through accidental wastewater discharge or direct discharge (Patel et al., 2021; Polo et al., 2020; Rimoldi et al., 2020).

The transmission of enteric viruses through recreational use of sewage-contaminated water being possible ((Lanrewaju et al., 2022; Wyn-Jones et al., 2011), the possibility and potential consequences of environmental contamination with SARS-CoV2 or other coronaviruses was raised early on.

Reports on the isolation of infectious SARS-CoV-2 from the feces and urine of COVID-19 patients have been documented but remain rare (reviewed in(Ahmed et al., 2021)). SARS-CoV-2 RNA was found to be significantly more persistent than infectious SARS-CoV-2, indicating that environmental detection of RNA alone does not substantiate the risk of infection and evidence of effective transmission is not clear (Bivins et al., 2020). SARS-CoV-2 transmission through wastewater and wastewater-contaminated waters is theoretically possible, but the actual risk is considered very low (Ahmed et al., 2021). Currently, an accurate risk assessment of viral exposure and transmission requires additional experimental evidence of CoV persistence in seawater.

Indeed, the resistance of virions to harsh conditions is highly variable and coronaviruses are considered fairly resistant in the environment, for review see (Adelodun et al., 2021; Silverman and Boehm, 2021). Indeed, SARS-CoV-2 virions, remained infectious for up to 3 h in aerosols and for 3 days on artificial surfaces respectively. SARS-CoV-2 virions are also stable over a wide range of pH values at room temperature (pH 3-10) (Anand et al., 2021; Chan et al., 2020; Chin et al., 2020; Kampf et al., 2020; Kwon et al., 2021; La Rosa et al., 2020a; La Rosa et al., 2020b; Nunez-Delgado, 2020). Previous studies on other CoVs (such as SARS-CoV, TGEV, and MHV) have shown them to be detectable in sewage for 2-4 days, in tap water for up to 10 days at 23– 25°C, and up to 100 days at 4°C, for review see (Giacobbo et al., 2021) but studies on the presence and persistence of CoVs in seawater remain scarce. One study showed that SARS-CoV-2 lost its infectivity within two days in seawater at 23°C (Lee et al., 2020), but half-lives were not calculated and this temperature is higher than those encountered in North-Western European countries like France. Another study calculated a T90 of 1.1 day in filtered seawater at 20°C for the porcine respiratory coronavirus (PRCV) (De Rijcke et al., 2021) and finally, a study showed a high impact of the temperature on the decay of SARS-CoV-2 in seawater by comparing conditions at 20°C and 4°C, with T90 of 7 to 10 days (Sala-Comorera et al., 2021). Intermediate temperatures were not tested. The possible long survival in water systems raises concerns about the persistence of SARS-CoV-2 or other CoV in the coastal environment.

Coronaviruses (order: Nidovirales, suborder: Cornidovirinae, family: Coronaviridae, subfamily: Orthocoronavirinae) show very strong genetic diversity and a high prevalence in nature (Ding and Liang, 2020). This family of viruses primarily infects a wide host spectrum of mammals and birds (Su et al., 2016). Based on the variety of non-structural accessory proteins, antigenic properties and host ranges, four genera of CoV have been described (Alpha-, Beta-, Delta-, and Gamma-CoV among which Alpha-and Beta-CoV include viruses infecting humans (HCoV). The emerged pandemic SARS-CoV-2 belongs to Beta-CoV, along with SARS-CoV-1 and MERS-CoV, two previously emerging HCoVs. Alpha and Betacoronaviruses are known to infect a variety of vertebrate mammalian hosts including swine, bats, minks or rodents. Until recently, deltacoronaviruses were known to infect a variety of hosts such as birds and non-human mammals, but two cases of human infection with porcine deltacoroanaviruses were reported in 2021 (Lednicky et al., 2021). Gammacoronaviruses infect essentially birds. Although coronaviruses are divided into four distinct genera, they all share similar physicochemical properties with an enveloped spherical viral particle of 120 nm diameter and the same genomic organization with a single-stranded RNA genome of positive polarity (25 - 30kb) coding for conserved ORFs that are the replicase complex, structural proteins (envelope, membrane, spike and nucleoprotein) and a variable number of non-structural accessory proteins. SARS-CoV-2, like some other coronaviruses (SARS-CoV, MERS) is zoonotic, leading to its classification as a BSL3-like pathogen with a commitment to need for an L3 laboratory facility for its manipulation. The number of BSL3 laboratory is much more limited than that of BSL2 laboratories and non-zoonotic coronaviruses have been largely and successfully used as surrogate of SARS-CoV-2 for studies in BSL2 facilities.

For this reason, we previously used porcine epidemic diarrhea virus (PEDV) and infectious bronchitis virus (IBV), both without any zoonotic potential, as viral substitutes for SARS-CoV-2 (Bernard louis et al., 2020) and found that treatment in a climate chamber at 70°C for 1 h with a 75% humidity rate was adequate for enabling substantial decontamination of both model animal coronaviruses, confirming for these two viruses the overall inactivation properties shared by coronaviruses (Guillier et al., 2020). In the current study, we relied on the CV777 strain of PEDv, a swine alphacoronavirus, as a surrogate for SARS-CoV-2.

To evaluate the stability and resistance of CoV and by extension of SARS-CoV-2 in the coastal environment, the genomic and infectious titers, the latter directly correlated to the infection risk of the CoV model and the SARS-CoV-2 surrogate, PEDv, were measured in natural coastal seawater for 28 days, and the results were used to build a mathematical model of coronavirus decay in seawater at a range of temperatures (8°C, 15°C, and 24°C) representative of the annual variation of the French coastal waters. A fourth temperature of 4°C was also investigated as a reference temperature, allowing for comparisons with other studies addressing decay in different types of water. The decay rate of PEDv was then used to determine the half-life of the virus in French coastal waters, using the temperatures reported for each trimester (year quarters) from 2000 to 2021.

## Materials and methods

### Cells and Virus

Vero cells (ATCC® CCL-81) are maintained in EMEM (ThermoFisher Scientific, France) supplemented with 10% fetal calf serum (reference 702774, Corning) and 1% Penicillin / Streptomycin (reference 11548876, GIBCO).

CV777 viral strain (genbank: AF353511) of porcine epidemic diarrhea virus (PEDV) was used as a surrogate of the SARS-CoV-2. Briefly, the virus is amplified on Vero cells, in an infection medium, composed of EMEM supplemented with 0.3% tryptone phosphate broth, 0.02% yeast extract, 1% Penicillin / Streptomycin and 10 μg / ml trypsin. (reference 215240, DIFCO). After 16 hours of infection, the cells are lysed by three successive freezing (−80 ° C), thawing (37 ° C). The culture medium is clarified by rapid centrifugation at 10000g for 10 minutes then the virus is pelletized for 4 hours at 20000g, and taken up in 1/100 of PBS of the initial volume of the CV777 inoculum. Viral titer was determined by Immuno Peroxidase Monolayer Assay (Detection limit: 0.5TCID50/ml), and genomic titer by one-step RT-qPCR (Detection limit: 50 copies/5µl of extract) (Bigault et al., 2020). The viral stock was titrated at 1.7*10e8 TCID50 / ml.

### Determination of the TCID50 by Immuno Peroxidase Monolayer Assay (IPMA)

In a 96-well plate, 8.10^4^ Vero cells are seeded per well and allowed to adhere for at least 6 hours. The cells are washed 3 times with PBS (sigma, France), then infected with 100 μl of virus diluted in the infection medium. The infection is stopped after 16 hours and the cells fixed with 50 µl of 80% acetone for 20 minutes at −20 ° C. After drying for 30 minutes at room temperature (RT), the endogenous peroxidazes are neutralized with 50 μl of a solution of 99% methanol / 1% H_2_O_2_ for 30 minutes at RT. Wells are washed twice with 200 μl of PBS tween (PBST, sigma # P3563), then blocked for 90 minutes at 37 ° C with a solution of PBST + 5% milk (PBST5, Dutscher # 2516188). The presence of viral proteins is detected by incubation with 100µl of an anti PEDV pig serum diluted to 1/300th in PBST5, 1h at 37 ° C, followed by 3 washes of 200µl PBST, then a 1h incubation at 37 ° C in the presence of 100µL of Goat anti pig IgG-coupled to horseradish peroxidase (HRP) (Sigma, France, #AP166P) diluted to 1/300 in PBST5. After three washes in PBST, the visualization is made by adding 50 μl of AEC/H_2_O_2_ (Reference AEC: sigma #A6926) as developer for 10 minutes RT. The reaction was stopped by removing the developer followed by a last wash with PBS. The viral titer was determined by the Kärber method (Kärber 1931). For analysis, log of TCID50 for each point were normalized as the ratio against the initial log of TCID50. TCID50 sensibility is as low as 0.5 * 10e1 TCID 50, which is therefore the low limit of detection.

### Viral decay trial

The impact of seawater temperature on the stability and survival of PEDv, chosen as a substitute model for SARS-CoV-2, is evaluated in natural coastal seawater collected and sand-filtered on the October 21^st^, 2020 at an experimental oyster farm on the French Atlantic coast (PMMB, Ifremer, Bouin, France) where it is used for growing oysters. At sampling, pH was 8.71, salinity 33.9 and turbidity 2.7 NTU. The seawater is then aliquoted and kept frozen at −80°C for 3 to 8 months before being used. The filtered coastal seawater was spiked with the CV777 virus stock to achieve a load of 1.10^6^ TCID50 / ml and 1.10^8^ genome copy / ml. To prevent any temperature variations due to sample manipulation, as well as the risk of contamination, 1 ml aliquots of this spiked water were incubated in water baths at 4 ° C, 8 ° C, 15 ° C and 24 ° C, in the dark, throughout the experiments. A preliminary experiment lasted 16 days with aliquots sampled randomly on day 0 to day 3, day 7 to day 10 and day 14 to day 16. Then, based on these first results three series of experiments were conducted during 28 days with aliquots randomly sampled on day 4 to day 7, day 11 to day 14, day 18 to day 21 and day 25 to day 28. For each time/temperature pair studied, aliquots were analyzed by PEDV-specific RT-qPCR, to quantify the viral genome, and by TCID50, to determine the infectious capacity of the virus. In addition, the rest of spiked seawater at day 0 was stored at −80 °C for later genomic load determination (Bigault 2020) and TCID50.

### Viral decay modeling

The objective here is to define a mathematical model representing the decrease in PEDv viral titers over time as a function of temperature. Two models were selected among those recently used for this type of analysis, a bi-exponential model and a Weibull type model (de Oliveira et al., 2021). Data were normalized for TCID 50, each point (_n_TCID50) treated as the ratio between the value obtained at point tn and the initial value at point t0.

#### - The bi-exponential model

This model is expressed as a function of 4 parameters: two threshold values a_1 and a_2 and two decay rates δ_1 and δ_2:

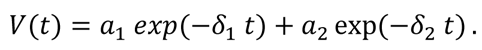

Thus, the initial titer is expressed by V(0)=a_1+a_2, to which two successive exponential decreasing phases.

#### - The Weibull model

### This model is also based on 4 parameters

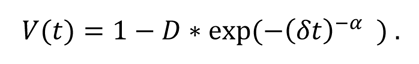

In this model, an asymptotic decrease is represented towards a threshold value *V*_oo_ = 1 - *D*. The parameter δ governs the initial phase of decay when the parameter α influences the behavior over longer durations and the speed of convergence towards the value *V*_oo_.

#### - Parameter estimation

The data used to estimate the parameters are shown in Figure 1 and Table 1. Each panel corresponds to a water temperature (4, 8, 15 and 24°C); for each of them, six kinetics were considered (tree separate experiments analyzed on duplicate). The estimation of the parameters was carried out using a non-linear mixed-effect model for which the kinetics of the repetitions are considered as a longitudinal follow-up in a population. Briefly, for each model, the parameters θ are estimated at the population level. The individual parameters were assumed to be log-normally distributed, thus ensuring their positivity. The parameters of individual i are given by

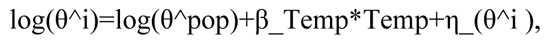

**Figure 1:**
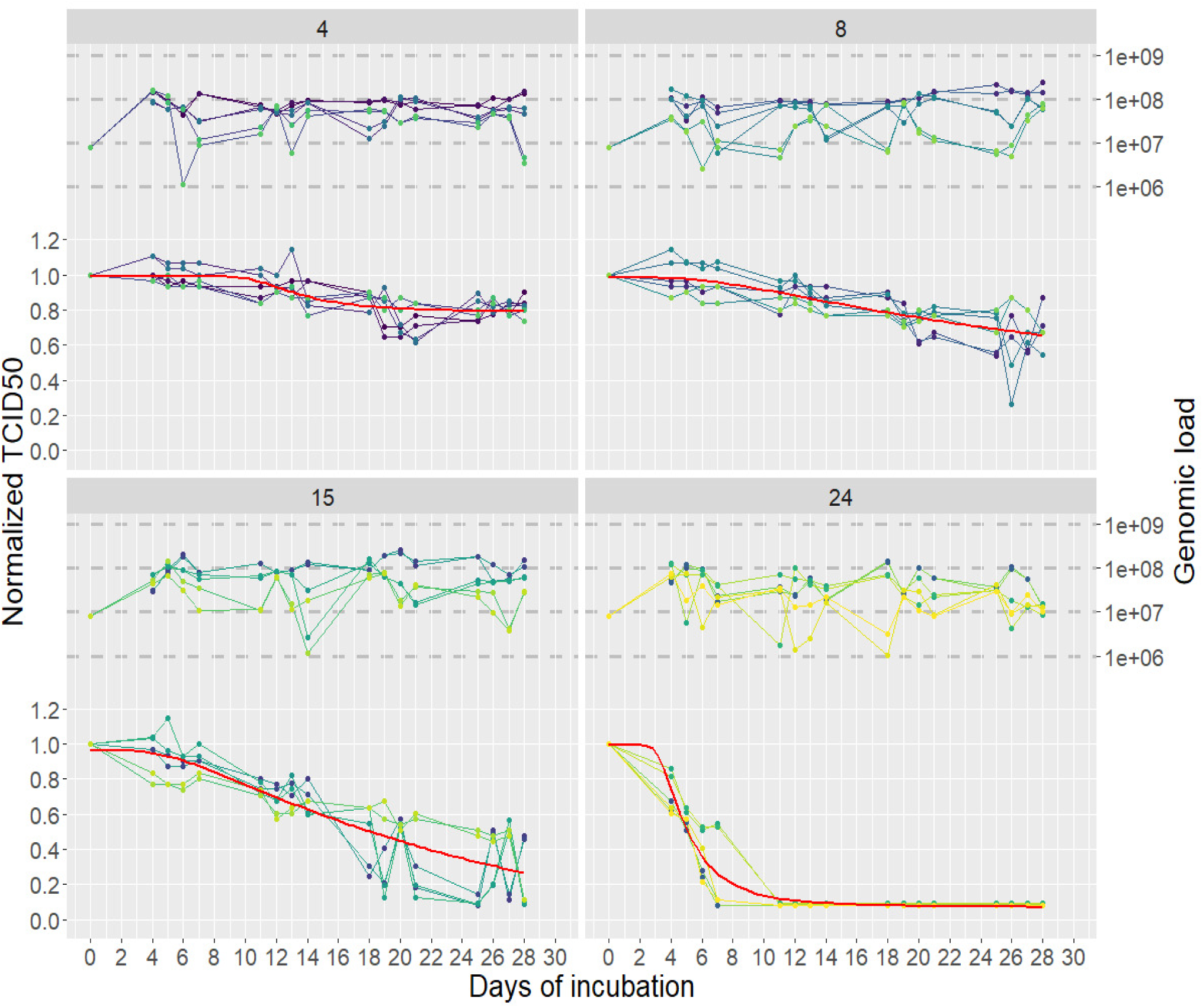
Genomic and infectious titers of PEDv incubated in seawater at four different temperatures for 28 days. 1 ml aliquots of seawater were spiked with the PEDv CV777 virus stock to achieve a load of 1.10^6^ TCID50 / ml and 1.10^8^ genome copy / ml then incubated in water baths for 28 days at 4 different temperatures : 4°C, upper left panel ; 8°C, upper right panel ; 15°C, lower left panel and 24°C, lower right panel. The genomic load (genome copies, in log scale, right axis, upper part of each panel) and infectious titer (ratio of the TCID50 measured at each time point (days of incubation, horizontal axis) on the one at day 0 for each experiment, left axis, bottom part of each panel) were measured 4 days a week (D4 to D7, D11 to D14, D18 to D21 and D25 to D28), by RT-qPCR and IPMA respectively, in duplicate for each aliquot on the same aliquot, and during three independent experiments. For each condition, the 6 measures are depicted with blue to yellow points and lines. The Weibull models calculated for the infectious titer decay are represented by the red curves.

**Table 1:** log 10 TCID50 and % from TCID50 at D0 of PEDv for viral titer quantification results during incubation in seawater at 4°C, 8°C, 18°C and 24°C, per days, during 28 days. 1 ml aliquots of water was spiked with the CV777 virus stock to achieve a load of 1.10e6 TCID50 / ml (6 log 10 TCID50) then incubated in water baths at the 4 different temperatures, for 28 days. Infectious titer were determined at day 0, 4 to 7, day 11 to 14 day 18 to 21 and day 25 to 28 by IPMA. **DPI: day post infection; SD: standard deviation; Temp: temperature.**

| Temp<br>DPI | Log10 TCID50 – Standard deviation |  |  |  |  |  |  |  |  |  |  |  |
| --- | --- | --- | --- | --- | --- | --- | --- | --- | --- | --- | --- | --- |
|  | 4 °C |  |  | 8 °C |  |  | 15 °C |  |  | 24 °C |  |  |
|  | mean | % | SD | mean | % | SD | mean | % | SD | mean | % | SD |
| 0 | 5.97 | 100.00 | 0.30 | 5.97 | 100.00 | 0.30 | 5.99 | 100 | 0.28 | 5.94 | 100 | 0.33 |
| 4 | 6.10 | 102.18 | 0.18 | 5.80 | 97.15 | 0.41 | 5.57 | 92.98 | 0.55 | 4.17 | 70.20 | 0.45 |
| 5 | 5.90 | 98.83 | 0.22 | 5.80 | 97.15 | 0.24 | 5.40 | 90.15 | 0.62 | 3.43 | 57.74 | 0.16 |
| 6 | 5.83 | 97.65 | 0.16 | 5.67 | 94.97 | 0.34 | 5.10 | 85.14 | 0.42 | 2.13 | 35.85 | 0.72 |
| 7 | 5.80 | 97.15 | 0.21 | 5.70 | 95.48 | 0.31 | 5.33 | 88.98 | 0.29 | 1.43 | 24.07 | 1.22 |
| 11 | 5.47 | 91.62 | 0.29 | 5.20 | 87.10 | 0.24 | 4.47 | 74.62 | 0.29 | 0.50 | 8.41 | 0.00 |
| 12 | 5.53 | 92.63 | 0.15 | 5.50 | 92.13 | 0.31 | 4.03 | 67.27 | 0.53 | 0.50 | 8.41 | 0.00 |
| 13 | 5.67 | 94.97 | 0.39 | 5.23 | 87.60 | 0.30 | 4.27 | 71.28 | 0.46 | 0.50 | 8.41 | 0.00 |
| 14 | 5.23 | 87.60 | 0.64 | 5.00 | 83.75 | 0.47 | 4.07 | 67.94 | 0.60 | 0.50 | 8.41 | 0.00 |
| 18 | 5.20 | 87.10 | 0.41 | 5.00 | 83.75 | 0.43 | 2.97 | 49.58 | 1.03 | 0.50 | 8.41 | 0.00 |
| 19 | 4.77 | 79.90 | 0.50 | 4.50 | 75.38 | 0.36 | 2.20 | 36.72 | 1.39 | 0.50 | 8.41 | 0.00 |
| 20 | 4.40 | 73.70 | 0.59 | 4.27 | 71.52 | 0.43 | 3.20 | 53.42 | 0.21 | 0.50 | 8.41 | 0.00 |
| 21 | 4.40 | 73.70 | 0.73 | 4.43 | 74.20 | 0.27 | 2.00 | 33.38 | 1.30 | 0.50 | 8.41 | 0.00 |
| 25 | 4.77 | 79.90 | 0.21 | 4.07 | 68.17 | 0.59 | 1.40 | 23.37 | 1.25 | 0.50 | 8.41 | 0.00 |
| 26 | 4.93 | 82.58 | 0.37 | 3.93 | 65.83 | 1.54 | 2.33 | 38.89 | 0.97 | 0.50 | 8.41 | 0.00 |
| 27 | 4.83 | 80.90 | 0.24 | 4.00 | 67.00 | 0.70 | 2.27 | 37.89 | 1.14 | 0.50 | 8.41 | 0.00 |
| 28 | 4.90 | 82.08 | 0.42 | 4.13 | 69.18 | 1.37 | 1.19 | 22.87 | 0.50 | 0.00 | 0.00 | 0.00 |

where θ_pop represents the median parameter independent of temperature at the population level. θ^pop represents the value of the mean parameter in the population. η_(θ^i) are random effects vectors assumed to be independent centered Gaussian vectors with variance ω_θ^2, representing interindividual variability. The temperature is integrated as a covariate for all the parameters and has an impact when the associated parameter β_Temp is significantly different from 0. The quality of adjustment of the models is evaluated according to the Akaike criterion (AIC), the model having the lowest AIC being selected.

#### - Seawater temperature along the French Coast

The temperature of the water is regularly monitored on different site of the seashore in France. In this study, data collected in continental French territory were considered. 308 sampling points were included for which temperature was recorded between 2000 and 2021 at different depth (REPHY dataset - French Observation and Monitoring program for Phytoplankton and Hydrology in coastal waters. Metropolitan data - https://doi.org/10.17882/47248). Here, we focused on surface waters only, with a depth varying between 0 and 1m. The dataset consisted in 54000 temperature records, which where analyzed quarterly from January to December to derive the average quarter temperature for each year. Feeding the decay model with these temperature records allowed for evaluating the virus half-life for each sampling location and each quarter.

## Results

### Survey of PEDv decay in seawater

The half-life of the coronavirus virion in seawater was poorly characterized when starting these experiments. Therefore, in a preliminary experiment (data not show), we measured the decay of virions at four different temperatures (4, 8, 15, and 24°C) for 16 days. At the two lowest temperatures, we observed only a slight decrease in infection of 5% and 12% at 4°C and 8°C, respectively, by day 16. At day 16, for the 15°C temperature the infectious titer dropped by about 40% (from 6 Log 10 TCID50 to 3,6 log 10 TCID50) and complete loss of infection was observed after 7 days at 24°C (0.8 log 10 TCID50). In parallel with infectivity, a measure of genomic load was performed using RT-qPCR, which revealed the stability of the viral genome throughout the 16 days duration of this preliminary experiment. Therefore, in order to be able to decipher more precisely the viral decay of PEDv in seawater, three independent series of experiments were performed with the same temperature settings (4, 8, 15 and 24°C) but extended to 28 days.

TABLE 1 gives the mean TCID50 results of the 3 series of experiments and Figure 1 shows the daily evolution of TCID50, normalized to the initial (D0) TCID50, for 28 days at the four different temperatures.

We observed good repeatability for the samples collected at different time points in the experiment, with the same trend for each of the triplicates according to the temperature. However, significant variability between the triplicates on days 20 to 28 at a temperature of 15 °C was also observed (day 25: 0.5 Log 10 TCID50 for series 1 and 2 and 2.9 Log 10 TCID50 for series 3), which gives a random and blurred aspect to this region of the curve. The complete experiment, over four weeks, confirmed the trend observed during the first test.

At 4°C and 8°C, we observed an overall good stability of the PEDv infectious titer with 82% and 69% of the initial TCID50 value maintained after 28 days of incubation in our coastal water sample, respectively. At 15°C, 88% of the TCID 50 was maintained during the first 7 days, and then the infectious titer dropped to 67% of the initial TCID50 during the next seven days followed by a regular decline of the TCID50 value to 23% of the initial TCID50 by day 28. In the last part of the experiment, the individual TCID50 values at 15°C were much more dispersed, as reflected by the increase in the standard deviation values for the last points from Day 18 onward. Finally, the complete loss of infectious PEDV after 7 days at 24°C was confirmed. In parallel with the measurement of the infectious titer, the viral genome concentration was measured by RT-qPCR to ascertain the presence of viral particles in the incubation medium used for the infection (Supplemental 1 and Figure 1). Despite day-to-day variations, on the 28 days duration of the experiment, the PEDv genome loads remained stable for the four experimental conditions, confirming that the decrease in infectious titer was not due to the loss of virus when preparing the viral inoculum for TCID50 experiments, but to the time-and temperature-dependent degradation of the viral particles.

### Modelling the infectious load decay of coronavirus in coastal water

Two models, a bi-exponential model and a Weibull type model (Camillo de Oliviera 2021) were evaluated to define a mathematical model of the decrease in PEDv viral titers in coastal water over time as a function of temperature. Akaike’s information criterion was lower with the Weibull model than with the bi-exponential model (−728 and −634, respectively), indicating a better fit of the data with the Weibull model. The model parameters, representing the average kinetics estimated at the population level are listed in Table 2.

**Table 2:** parameters estimated at the population level representing the average kinetics for the bi-exponential and the Weibull model. **(* p-value<0.05).** AIC : Akaike’s information criterion.

|  | Bi-exponential model parameters |  |  |  | Weibul model |  |  |
| --- | --- | --- | --- | --- | --- | --- | --- |
| | $a_1$ | $\delta_1$ | $a_2$ | $\delta_2$ | $D$ | $\delta$ | $\alpha$ |
| Fixed effect | 0.88 (0.11) | 0.003<br>(0.001) | 0.16 (0.02) | 0.06 (0.08) | 0.71 (0.09) | 0.02 ( 0.002) | 0.48 (0.09) |
| Temperature<br>Covariate | 0.006 (0.005) | 0.17* (0.02) | -0.03 (0.05) | -0.03 (0.11) | 0.01* (0.005) | 0.1* (0.007) | 0.08* (0.01) |
| AIC | -638 |  |  |  | -737 |  |  |

The log-linear regression model revealed a significant effect of temperature on all the individual parameters, accounting for inter-individual variability. Correlation between the parameters estimated for each kinetic and the water temperature is shown in Figure 2. The parameters governing the decrease (δ and α) reflect the persistence observed at 4°C over the duration of the experiment. These parameters showed a strong dependence on the water temperature. A strong exponential increase in δ was observed between 15 °C and 24°C, indicating an impact on the initial decrease. The increase in parameter α, which governs the speed of convergence towards V_∞ (asymptotic viral load), also showed an exponential trend, varying from 1 to 3 between 4 and 24°C.

**Figure 2:**
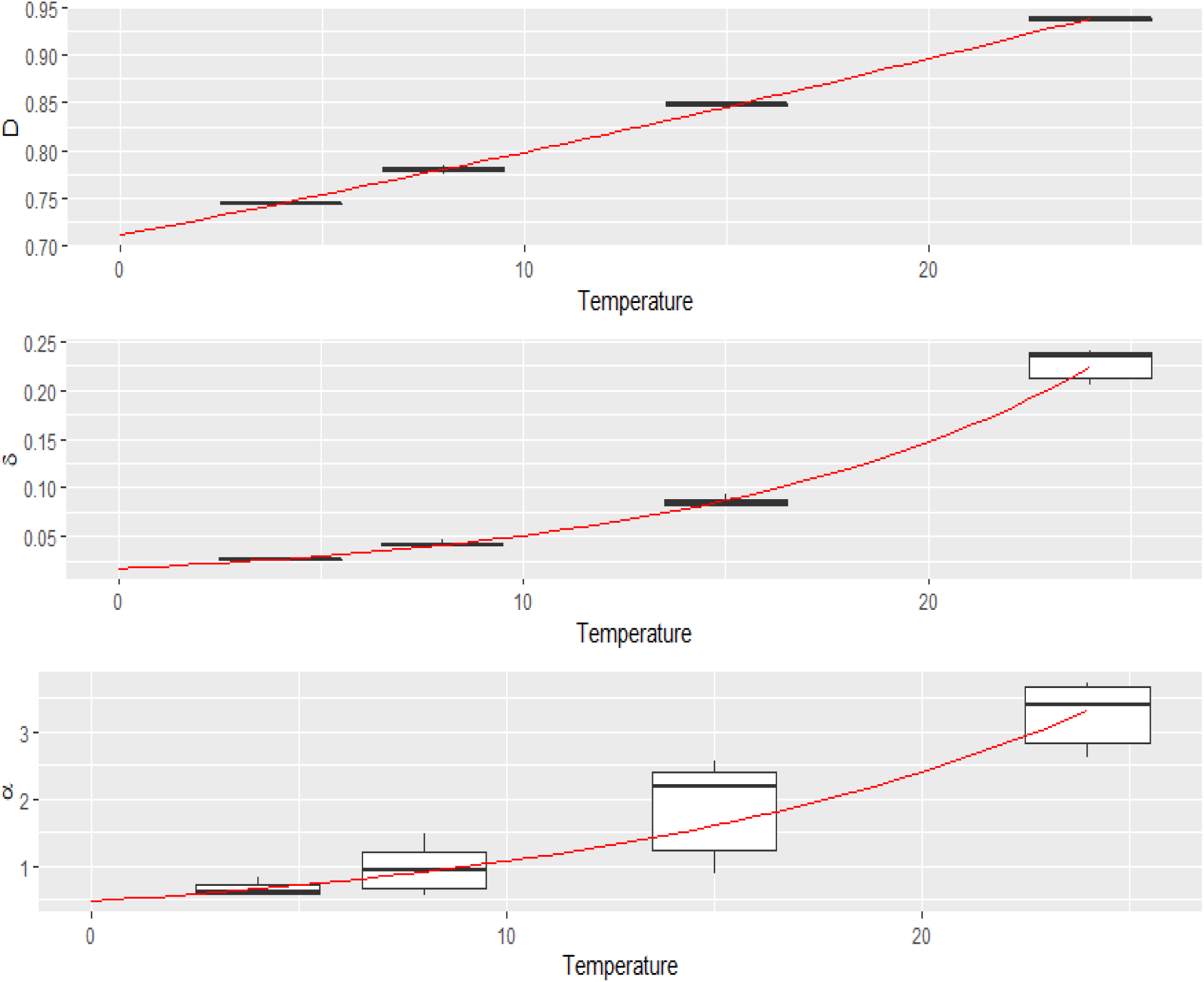
Correlation between the parameters estimated for each kinetic and the water temperature.

### Analysis of the persistency of coronaviruses along the French seashore

We applied the coronavirus decay model along the French seashore, considering the variations in temperature occurring within a year.

The quarterly average temperature varied seasonally, ranging from 6 °C to 14°C in winter and from 14 to 26°C in summer. The map displayed in figure 3 highlights geographical discrepancies between northern and southern France as well as between the Mediterranean coast and the Atlantic Ocean in the west.

**Figure 3:**
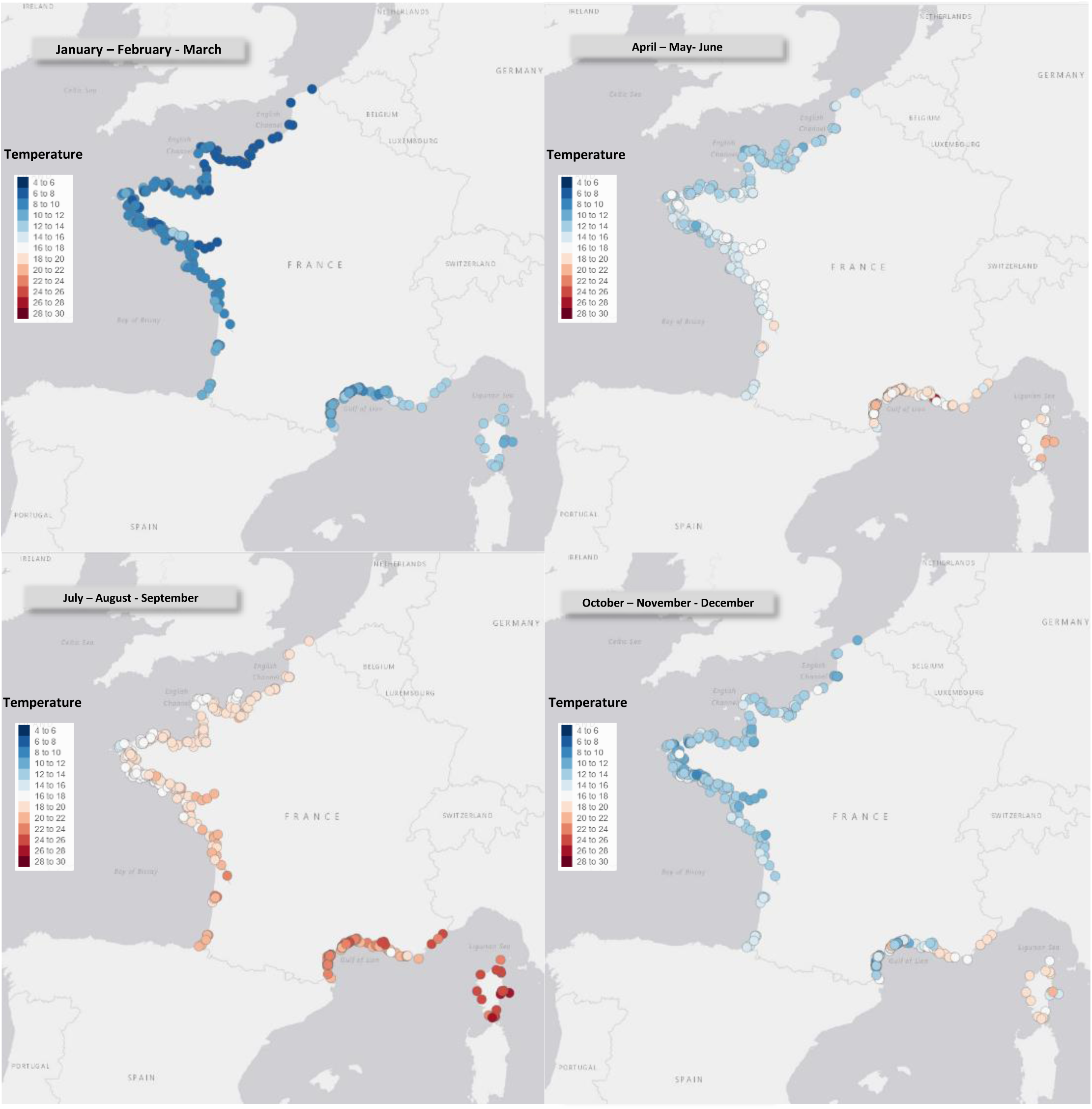
Mean quarterly temperatures of surface waters on the French coast from 2000 to 2021.

The half-life of the virus in the surface water at each sampling point was evaluated based on the quarterly average temperature and is represented as a heat map in figure 4. Reflecting the analysis of the temperature records, a huge seasonal effect was observed, with a half-life of coronavirus infectivity of up to 70-80 days during the coldest period, falling below 20 days at all locations during the summer. The northern area exhibited the highest half-life seasonal variation compared to that of Corsica where water temperatures remained much stable throughout the year.

**Figure 4:**
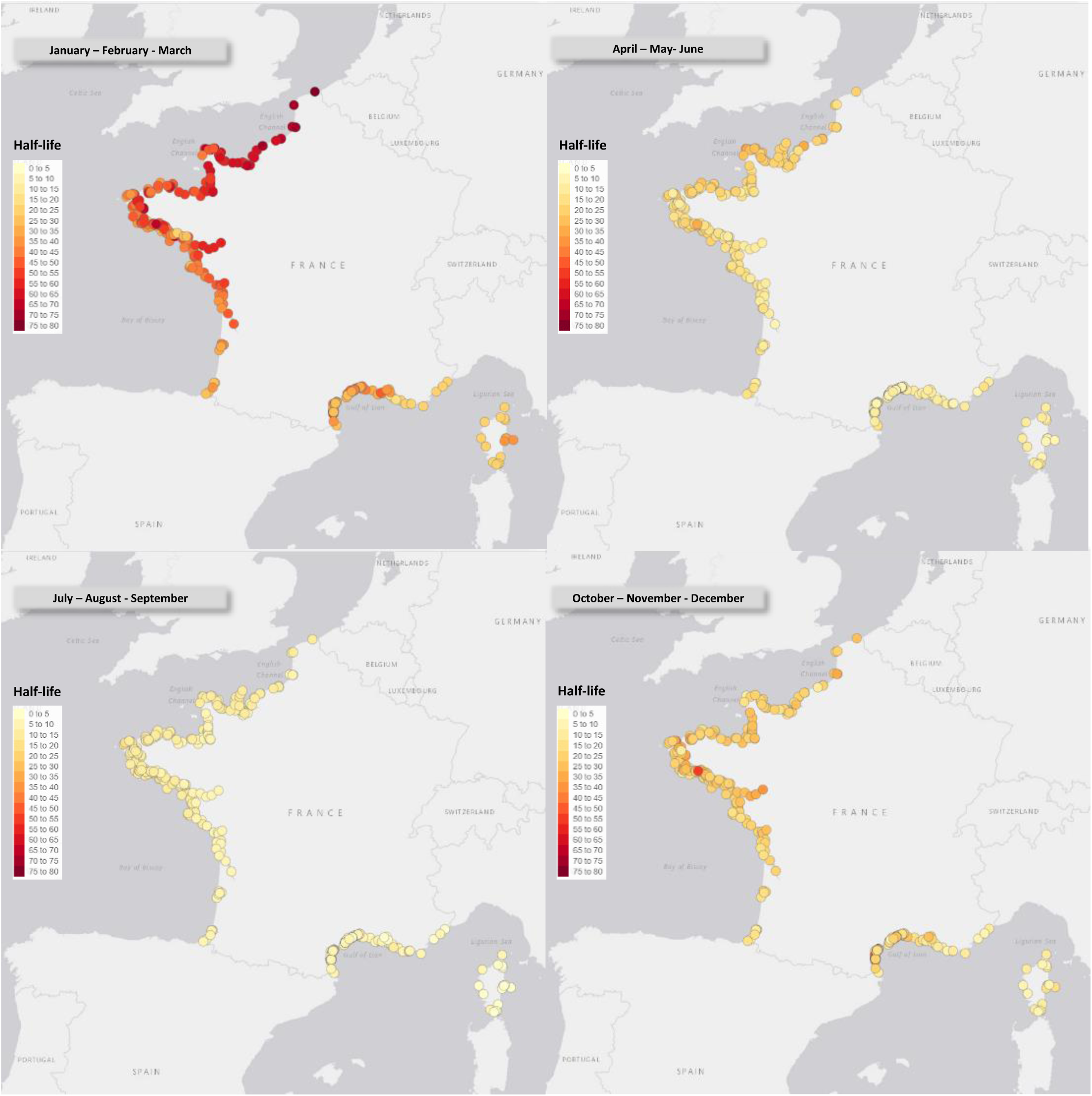
Estimated half life (in days) of coronavirus in surface water, based on quarterly average temperature for each collection location.

## Discussion

Viral contamination of water or food by human sewage is a long-documented origin of gastroenteritis outbreaks, as exemplified by the contamination of oysters by human norovirus (HuNoV) (Le Guyader et al., 2009; McLeod et al., 2017). At the beginning of the pandemie, the question regarding SARS-CoV-2 risk of exposure and transmission to human during recreational activities was highlighted. Although significant amounts of SARS-CoV-2 RNA are regularly detected in the feces of patients, it has been shown that the infectious capacity of the SARS-CoV-2 virus from feces is very limited. A few studies have concluded that the risk of infection following exposure to SARS-CoV-2 in water is low (Ahmed et al., 2021; Bivins et al., 2020; Silverman and Boehm, 2021). However, the evaluation of CoV survival in water and especially in seawater will be informative for SARS-CoV-2 but also for other CoV, especially enteric CoV.

This study aimed to monitor experimentally and concretely the survival/persistence of PEDV, as a representative of CoV and as a surrogate for SARS-CoV-2, in marine waters. This using the modeling of its decay over time and water temperature data taken in France. The potential risk of infection in French coastal waters, in case of contamination by the discharge of wastewater was also evaluated and confirmed to be low or inexistent. As mentioned above, SARS-CoV-2 manipulation requires a level 3 laboratory, which is available in a limited number of facilities and represents more tedious working conditions than level two laboratories. To circumvent this constraint, several surrogate agents of SARS-CoV-2 have been used for decay and environmental persistence experiments. Non-enveloped viruses, such as bacteriophage MS2, pose problems of misinterpretation and accuracy; non enveloped viruses are known to be more resistant than enveloped viruses (La Rosa et al., 2020a; La Rosa et al., 2020b). The use of a coronavirus as a surrogate provides greater reliability as coronaviruses form an organizational unit with similar biophysical properties and genomic structures. Moreover, (Guillier et al., 2020) have shown a similar temperature and relative humidity persistence potential of different coronaviruses infecting human and non-human mammals, on surfaces and in suspension, confirming the potential of animal coronaviruses as surrogates. To study the persistence of coronavirus in seawater, we selected PEDv, a porcine enteric alphacoronavirus we handled in a BSL2-type confined laboratory. Recently, the PEDv has been used several times, as surrogate for HCoV, including SARS-CoV-2, i) to compare analytical methods to detect SARS-CoV-2 in wastewater (Perez-Cataluna et al., 2021), ii) as a process control for the monitoring of the occurrence of SARS-CoV-2 in 2020 in six wastewater treatments plants in Spain (Randazzo et al., 2020), iii) to evaluate the use of viability RT-qPCR for the selective discrimination and surveillance of infectious SARS-CoV-2 in secondary-treated wastewater (Monteiro et al., 2022), iv) to test the temperature sensitivity of different CoV in fomites (Cuevas-Ferrando et al., 2022), v) to evaluate the effects of dry-and moist-air controlled heating treatment on structure and chemical integrity, decontamination yield, and filtration performance of surgical masks and FFP2 respirators (Bernard et al., 2020), vi) and to test inactivation of PEDv on contaminated surgery masks by low-concentrated sodium hypochlorite dispersion (Antas et al., 2020). Furthermore, in a previous study, we demonstrated and quantified the bioaccumulation of PEDv or inactivated SARS-CoV-2 in oysters, and observed a similar tissue distribution and efficacy of bioaccumulation between the two coronaviruses, suggesting that PEDv is an adequate surrogate for SARS-CoV-2 in marine environments (Desdouits et al., 2021).

Various studies using surrogate viruses (Sala-Comorera et al., 2021) and addressing the detection and survival of coronaviruses in wastewater, river water, or filtered or sterilized water have been conducted over the past 2 years (Ahmed et al., 2020b; Guillier et al., 2020; Randazzo et al., 2020). Others used both surrogates and the SARS-CoV-2 (de Oliveira et al., 2021; Sala-Comorera et al., 2021) but were conducted either at low temperatures (4°C) or at higher temperatures (20–25°C). Yet, the temperature gradient was not considered in the same model. The synthesis that can be drawn from these studies shows a common trend, in which different viruses remain infectious for several hours to several weeks, depending on the experiment, in different types of water (pure water, surface water, seawater and sterilized wastewater), with infectivity maintained for a longer period of time at the lowest temperatures (Chin et al., 2020; Lee et al., 2020; Ye et al., 2016). Recently, (Sun et al., 2022) showed an inverse correlation between temperature and viral titer for the viability of SARS-CoV-2 in media and artificial seawater. These studies illustrate that intact coronavirus particles present in coastal waters may remain infectious for some time.

Our results present a follow-up of the decay of the virus over a long period with a wide temperature gradient (from 4°C to 24°C), similar to the temperature gradient observed in French coastal waters throughout the year, associated with a mathematical model used for evaluating the virus half-life along the French shore throughout the year. This experiment clearly shows that seawater temperature has a dramatic effect on the duration of the infectious capacity of PEDv and, by homology, on coronaviruses including SARS-CoV-2. The parallel evaluation of the genomic load of the samples throughout the experiment precludes the possible bias of a reduction in the viral load due to artifacts such as adsorption on the tube walls or other phenomena. Under summer conditions representative of Mediterranean conditions (24°C), we observed a rapid loss of the infectious capacity of the virus in seawater, with a loss of almost two log TCID50 at 3-4 days and a total loss at day 7. Under conditions mimicking the spring Atlantic coast (∼15°C), the effect was more gradual, with a loss of one log in one week and almost total loss at 3 weeks. In winter conditions (4°C and 8°C), the infectious virus is more stable, with survival extending beyond four weeks

In another study assessing SARS-CoV-2 survival in cell culture medium, with a similar range of temperatures as ours (4°C, 13°C, 21°C and 25°C), the best virus survival was also observed with the coolest conditions; the virus was relatively stable for all the temperature conditions with the maximum log reduction of 1.17 virus titer at 4 days post-contamination at 25°C (5.104 TCID50 starting dose) (Kwon et al., 2021). Coronaviruses are sensitive to extremely acidic and basic pH. Therefore, the faster inactivation of PEDv in seawater can be explained by the high salinity concentration in combination with alkaline conditions (pH>8) in seawater compared to that in culture medium (Kwon et al., 2021), wastewater and pure water (Chan et al., 2020).

Using the experimental kinetics of the viral decay of PEDv as a function of real temperature data along the French coast during the year (considering the mean temperatures reported from 2000 to 2021) allowed us to evaluate the half-life of the virus according to seasons. The risk of viral transmission is correlated with a longer half-life of the virus and a higher frequentation of human populations in coastal environments. Fortunately, these two parameters are inversely correlated with a peak of human population in coastal areas during summer, when the half-life of the virus is the shortest, owing to the higher water temperature which is deleterious to the virus. Conversely, the better survival conditions for the virus in winter is balanced by a lower population size and frequentation in the recreational usage of coastal areas. The situation is somewhat different concerning workers, such as for oyster farmers or fisherman, in contact with water all the year around even at the coldest temperature when virus half-life is the longest. In our previous study, we showed a low bioaccumulation efficiency of SARS-CoV-2 in oysters, as well as the absence of detection of this virus in seawater and shellfish samples collected on the French coast from April to August 2020 (Desdouits et al., 2021), confirming that the actual risk of contamination in food such as shellfish by the SARS-CoV-2 is low. Furthermore, if we consider the effect of tides, bringing an additional physical dispersion and dilution effect on the virus, all of these parameters favor a low risk of coronavirus contamination during seashore activities.

It is important to mention that these results could be refined by considering additional parameters of seawater as the variation in salinity (this study used only one salinity representative of the Brittany coast) and by estimating the effect of solar radiation, a known driver of viral decay, with summer exhibiting higher UV radiation exposure than winter.

Given the low survival of SARS-CoV-2 in sewage (Silverman and Boehm, 2021), the main risk of SARS-CoV-2 contamination through coastal water exposure likely lies in the direct release of raw sewage, places without connection to wastewater collection systems, ports, and in the event of a sanitation accident or human feces in seawater. The question also arises of soils contamination during spreading by slurry potentially contaminated by coronaviruses from livestock (ex: PEDv) and therefore by runoff and water contamination. This work could thus be used for the identification of areas that are the most at risk for humans in the case of spillover before treatment, as well as for the identification of maritime areas at risk of reverse zoonotic transmission to the marine animal population and, therefore, to be monitored as a priority. Indeed, cases of SARS-CoV-2 contamination have been reported essentially in various terrestrial animal species (Andersen et al., 2020; Hale et al., 2021; Prince et al., 2021; Sharun et al., 2021) it might also infect marine mammals (Audino et al., 2021). Alpha, and gammacoronavirus infections are already known to cause respiratory diseases in aquatic mammals and recent studies have shown that several species of marine mammals possessed the SARS-CoV-2 receptor, ACE2 (Luan et al., 2020; Nabi and Khan, 2020), with amino acids sequences highly conserved between human and marine mammal species. The binding of SARS-CoV-2 to ACE2 is an essential step in the infection of SARS-CoV-2, which can therefore make these animals susceptible to SARS-CoV-2 infection (Mathavarajah et al., 2021; Mordecai and Hewson, 2020). Thus, contamination of seawater from sewage, wastewater effluent, or urban and agricultural runoff, and the survival of SARS-CoV-2 are potentially a risk for marine mammals. Guo et al. modeled this risk by analyzing the possible dispersion of the virus following contamination in seawater and confirmed that this risk of infection is directly linked to viral concentrations, but also suggested a critical role of the temperature of the water (Guo et al., 2021).

## Data Availability

All data produced in the present study are available upon reasonable request to the authors

