## Supplemental S1 for "Evaluation of coronavirus decay in French coastal water and application to SARS-CoV-2 risk evaluation using Porcine Epidemic Diarrhea Virus as surrogate"

**Supplementary Material**

**Figure S1: Viral genomic load (copies /ml) results for the survey of PEDv in seawater at 4°C, 8°C, 18°C and 24°C, per days, during 28 days.**

**Figure legends**

|  | **Viral genomic load (copies /ml) - Standard deviation** | | | | | | | |
| --- | --- | --- | --- | --- | --- | --- | --- | --- |
| **Temp** | 4 °C | | 8 °C | | 15 °C | | 24 °C | |
| **DPI** | mean | SD | mean | SD | mean | SD | mean | SD |
| 4 | 1.31E+08 | 3.37E+07 | 9.24E+07 | 4.99E+07 | 5.04E+07 | 1.86E+07 | 8.25E+07 | 3.46E+07 |
| 5 | 8.45E+07 | 2.23E+07 | 5.10E+07 | 3.89E+07 | 1.00E+08 | 2.69E+07 | 6.47E+07 | 4.45E+07 |
| 6 | 4.88E+07 | 2.48E+07 | 6.76E+07 | 4.29E+07 | 1.08E+08 | 7.06E+07 | 6.53E+07 | 3.65E+07 |
| 7 | 5.88E+07 | 5.89E+07 | 2.79E+07 | 2.54E+07 | 5.62E+07 | 2.82E+07 | 2.64E+07 | 1.16E+07 |
| 11 | 5.04E+07 | 2.45E+07 | 5.61E+07 | 3.99E+07 | 6.86E+07 | 5.31E+07 | 3.47E+07 | 2.26E+07 |
| 12 | 5.63E+07 | 8.85E+06 | 6.16E+07 | 2.93E+07 | 7.63E+07 | 1.16E+07 | 3.65E+07 | 3.64E+07 |
| 13 | 4.82E+07 | 2.94E+07 | 6.31E+07 | 2.32E+07 | 5.82E+07 | 3.56E+07 | 3.75E+07 | 2.35E+07 |
| 14 | 7.49E+07 | 2.16E+07 | 4.60E+07 | 3.26E+07 | 5.12E+07 | 5.94E+07 | 2.48E+07 | 9.06E+06 |
| 18 | 5.32E+07 | 3.11E+07 | 5.41E+07 | 3.74E+07 | 1.01E+08 | 3.79E+07 | 7.02E+07 | 6.09E+07 |
| 19 | 6.01E+07 | 3.15E+07 | 7.52E+07 | 2.46E+07 | 1.11E+08 | 6.35E+07 | 2.63E+07 | 4.28E+06 |
| 20 | 7.21E+07 | 3.52E+07 | 7.68E+07 | 4.79E+07 | 1.01E+08 | 1.10E+08 | 5.32E+07 | 4.15E+07 |
| 21 | 6.97E+07 | 2.78E+07 | 8.91E+07 | 6.17E+07 | 6.19E+07 | 5.56E+07 | 3.04E+07 | 2.36E+07 |
| 25 | 4.50E+07 | 2.17E+07 | 7.78E+07 | 8.30E+07 | 8.73E+07 | 7.67E+07 | 3.35E+07 | 5.42E+06 |
| 26 | 6.04E+07 | 2.20E+07 | 6.24E+07 | 7.25E+07 | 5.10E+07 | 3.67E+07 | 4.05E+07 | 4.68E+07 |
| 27 | 6.78E+07 | 2.86E+07 | 9.21E+07 | 4.38E+07 | 3.97E+07 | 2.85E+07 | 2.97E+07 | 2.12E+07 |
| 28 | 6.76E+07 | 6.34E+07 | 1.09E+08 | 7.24E+07 | 7.29E+07 | 4.84E+07 | 1.22E+07 | 2.42E+06 |

**Figure S1: Viral genomic load (copies /ml) results for the survey of PEDv in seawater at 4°C, 8°C, 18°C and 24°C, per days, during 28 days.** 1 ml aliquots of water was spiked with the CV777 virus stock to achieve a viral genomic load of 10E8 then incubated in water baths at the 4 different temperatures, for 28 days. Infectious titer were determined at day 0, 4 to 7, day 11 to 14 day 18 to 21 and day 25 to 28 by one step RT-qPCR**. DPI: day post infection; SD: standard deviation; Temp: temperature.**
